# Low Dose Radiation Therapy for COVID-19 Pneumonia: A Pilot Study

**DOI:** 10.1101/2020.11.16.20231514

**Authors:** Daya Nand Sharma, Randeep Guleria, Naveet Wig, Anant Mohan, Goura Kisor Rath, Vellaiyan Subramani, Sushma Bhatnagar, Supriya Mallick, Aman Sharma, Pritee Patil, Karan Madan, Manish Soneja, Sanjay Thulkar, Angel Rajan Singh, Sheetal Singh

**Affiliations:** Department of Radiation Oncology, All India Institute of Medical Sciences, New Delhi 110029 India; Department of Pulmonary Medicine, All India Institute of Medical Sciences, New Delhi 110029 India; Department of Medicine, All India Institute of Medical Sciences, New Delhi 110029 India; Department of Onco-anesthesia and Palliative Medicine, All India Institute of Medical Sciences, New Delhi 110029 India; Department of Radiology, All India Institute of Medical Sciences, New Delhi 110029 India; Department of Hospital Administration, All India Institute of Medical Sciences, New Delhi 110029 India

**Keywords:** COVID-19, pneumonia, radiation therapy, low dose

## Abstract

**Background:** The World Health Organization (WHO) has declared coronavirus disease 2019 (COVID-19) as pandemic in March 2020. Currently there is no vaccine or specific effective treatment for COVID-19. The major cause of death in COVID-19 is severe pneumonia leading to respiratory failure. Radiation in low doses (<100 cGy) has been known for its anti-inflammatory effect and therefore, low dose radiation therapy (LDRT) to lungs can potentially mitigate the severity of pneumonia and reduce mortality. We conducted a pilot trial to study the feasibility and clinical efficacy of LDRT to lungs in the management of patients with COVID-19.

**Methods:** From June to Aug 2020, we enrolled 10 patients with COVID-19 having moderate to severe risk disease [National Early Warning Score (NEWS) of ≥5]. Patients were treated as per the standard COVID-19 management guidelines along with LDRT to both lungs with a dose of 70cGy in single fraction. Response assessment was done based on the clinical parameters using the NEWS.

**Results:** All patients completed the prescribed treatment. Nine patients had complete clinical recovery mostly within a period ranging from 3-7 days. One patient, who was a known hypertensive, showed clinical deterioration and died 24 days after LDRT. No patients showed the signs of acute radiation toxicity.

**Conclusion:** Results of our study (90% response rate) suggest the feasibility and clinical effectiveness of LDRT in COVID-19 patients having moderate to severe risk disease. This mandates a randomized controlled trial to establish the clinical efficacy of LDRT in COVID-19 pneumonia.

## INTRODUCTION

Currently, there is a global outbreak of COVID-19 caused by severe acute respiratory syndrome coronavirus 2 (SARS-CoV-2). On March 11, 2020, the WHO has declared COVID-19 as a pandemic and as on 14^th^ Novemebr 2020, there are a total of 52487476 confirmed cases and 1290653 deaths. **(1)** The clinical spectrum of COVID-19 varies from asymptomatic to clinical conditions characterized by respiratory failure that necessitates mechanical ventilation and intensive care unit (ICU) support. Severe pneumonia progressing to acute respiratory distress syndrome (ARDS) is the cause of death in majority of cases. The death rate ranges from 2-15% in different group of patients. **(2,3,4)**

Currently, there is no vaccine or specific antiviral drug available for COVID-19. The treatment is symptomatic, and oxygen therapy represents the major treatment intervention for patients with severe infection. Based on severity of symptoms, there are several scoring methods which can predict the outcome of disease. The NEWS is a popular scoring system used for non-ICU patients suffering from acute illness. **(5)** This score helps in quickly determining the degree of illness and intervention required. It also provides likelihood of death or admission to an ICU. Based on this score, any illness can be categorized as mild (Score <4), moderate (Score 5-6 or individual 3) and severe (Score ≥7). According to this scoring system, COVID-19 patients having a score of 5-6 will have probability of 15.7% critical events (ICU admission or 30-day mortality) and those having score≥7 will have about 24.1% critical events.**(5)**

The pathogenesis of COVID-19 pneumonitis appears to involve a cytokine storm with elevated pro-inflammatory cytokines such as IL-6 and TNFα among many others, leading to respiratory failure. **(6)** Radiation therapy (RT) in low doses (<100 cGy) is known to have its anti-inflammatory action by downregulating pro-inflammatory macrophages and upregulating anti-inflammatory macrophages (interleukin-10, transforming growth factor β1) and natural killer (NK) T cells thus countering the immune reaction incited by COVID-19. **(7)** Low dose lung irradiation can potentially mitigate the severity of pneumonitis thereby reducing the risk of death. It was a popular treatment of viral pneumonias until 1940s. **(8)** Historical data **(9-13)** suggests that LDRT to whole lung can possibly prevent cytokine storm and ARDS. A few recent trials **(14, 15)**, though with small sample size and short-term results have shown encouraging results with LDRT doses ranging from 0.5-1.5 Gy. We therefore conducted a pilot trial to study the feasibility and clinical efficacy of LDRT to lungs in the management of patients with COVID-19.

## METHODS

This pilot study was conducted at our Institute to assess feasibility and clinical efficacy of LDRT in patients with COVID-19. The study was initiated after approval by Institute Ethics committee (Ref. No. IEC-465/22.05.2020, RP-01/2020). It is registered on ClinicalTrials.gov (NCT04394793) and Clinical Trials Registry India (CTRI/2020/06/025862). The study protocol was designed jointly by the radiation oncology team and Institute COVID Management Team (comprising of members from departments of Internal medicine, pulmonary medicine, intensive care, hospital infection control committee). The sample size of 10 patients was determined based on multiple factors like incidence of COVID-19 disease, severity of disease, risk of viral exposure to radiation oncology team, availability of radiation therapy resources, distance between the COVID-19 indoor unit and radiation therapy machine etc. Eligible patients (as per inclusion criteria) were enrolled in the study after obtaining their consent. In order to avoid viral exposure to team, video conference facility was used while counseling them and taking their consent. Complete details about the trial, steps involved, benefits or side effects of treatment were explained to every patient.

### Eligibility Criteria

Eligibility criteria included age more than 18 years, diagnosis of COVID-19 confirmed by RT-PCR and moderate to severe illness (NEWS ≥5). Generally, the febrile patients who were already admitted to our indoor unit for COVID-19 management and having respiratory rate of >24 per minute and or oxygen saturation of <94% were screened for inclusion as they were likely to fulfill the eligibility criteria. Patients requiring mechanical ventilatory support or having unstable hemodynamic status were excluded. As COVID-19 virus is highly contagious and poses a risk to the various staff members during laboratory and radiological investigations, we adopted NEWS for inclusion of patients as well as response assessment as this is mostly based on clinical parameters **(Table 1)**. We targeted patients having moderate to severe disease to assess the efficacy of LDRT. The study outcome measures were: 1) number of ICU admissions or deaths; 2) improvement in NEWS score post LDRT and 3) length of hospital stay post LDRT.

**Table 1:** National Early Warning Score (NEWS) formula

| Criteria | Point Value |
| --- | --- |
| **Respiratory Rate (breaths per minute)** |  |
| ≤8 | +3 |
| 9-11 | +1 |
| 12-20 | 0 |
| 21-24 | +2 |
| ≥25 | +3 |
| **Oxygen Saturation (%)** |  |
| ≤91 | +3 |
| 92-93 | +2 |
| 94-95 | +1 |
| ≥96 | 0 |
| **Any Supplemental Oxygen** |  |
| Yes | +1 |
| No | 0 |
| **Temperature in °C (°F)** |  |
| ≤35.0 (95) | +3 |
| 35.1-36.0 (95.1-96.8) | +1 |
| 36.1-38.0 (96.9-100.4) | 0 |
| 38.1-39.0 (100.5-102.2) | +1 |
| ≥39.1 (≥102.3) | +2 |
| **Systolic BP** |  |
| ≤90 | +3 |
| 91-100 | +2 |
| 101-110 | +1 |
| 111-219 | 0 |
| ≥220 | +3 |
| **Heart Rate (beats per minute)** |  |
| ≤40 | +3 |
| 41-50 | +1 |
| 51-90 | 0 |
| 91-110 | +1 |
| 111-130 | +2 |
| ≥131 | +3 |
| **AVPU (Alert, Voice, Pain, Unresponsive)** |  |
| A | 0 |
| V, P, or U | +3 |

**Adapted from:** Royal College of Physicians. National Early Warning Score (NEWS): Standardizing the assessment of acute illness severity in the NHS. Report of a working party. London: RCP, 2012.

### Treatment and workflow to RT machine

All patients were treated as per the Institute COVID-19 standard management guidelines along with intervention of LDRT to both lungs with a dose of 70cGy in single fraction. The standard medical treatment generally consisted of oxygen supplementation, antibiotics, dexamethasone and general supportive care. LDRT was delivered employing 2 opposed antero-posterior and postero-anterior open portals. We did not use CT based RT planning as the prescribed dose is very low and the expected dose to organs at risk (OAR) is negligible. Additionally, CT simulation would increase the risk of viral exposure to staff. For patient transport from indoor unit to RT machine (Varian True Beam Radiotherapy System Linear Accelerator), the corridor was isolated. Patients were made to wear personal protective equipment (PPE), or mask covering face and head before they were shifted to RT area. The patient was positioned supine with hands above head. Machine gantry was rotated to 90 degree and a lateral chest X-ray portal image was taken in order to guide in measuring dose prescription point. Gantry was rotated back to 0 degree; an adequate field size was opened in order to cover both lungs. Upper border of the field was kept above the superior edge of lateral end of clavicle extending 1 cm superior to lung apex on portal imaging. Lower border extended below at the level of the L1 vertebrae (transpyloric plane) in order to cover costophrenic angle. Lateral borders were in the air on both sides of chest. A dose of 70 cGy in single fraction was prescribed at the midpoint of anterior-posterior chest separation using 6MV X-ray photons. Oxygen saturation level was continuously monitored during the entire treatment by placing the pulse oximetry monitor facing the CCTV camera. After completion of treatment, the patient was transported back to indoor unit through the same corridor. The Linear Accelerator unit was sanitized as per the cleaning and disinfection guidelines provided by the vendor.

### Response Assessment

The response assessment was done mainly on clinical parameters as per the NEWS. Imaging, routine hematological investigations and serum markers levels (like C-reactive peptide, D-dimer, interleukin-6, ferritin etc.) were done as and when needed, but not mandatorily for response assessment. The NEWS was recorded on Day 3, Day 7 and Day 14. These scores were compared with baseline score recorded on the day of LDRT i.e. on Day 0. Clinical response was defined as subject achieving NEWS of 0 within 14 days following LDRT. Failure was defined as ICU admission anytime after LDRT or death within 30 days. Patients were generally discharged from the hospital after they attained NEWS of 0 along with negative test for COVID-19. They were contacted on phone for any further required information.

## RESULTS

A total of 10 patients were recruited and treated from June to Aug 2020. **Table 2** shows various clinical characteristics of the patients. All the patients were male with a median age of 51 years (range 38 to 63 years). The shortness of breath was the commonest symptoms, found in all patients. The median respiratory rate was 22 per minute (range 21-27 per minute). Majority of patients (8 out of 10) had NEWS of 5-6. Three patients had co-morbidities (hypertension, 2 and diabetes, 1).

**Table 2:** Clinical Characteristics of the patients.

| Patient Serial No. | Age group (year) | Sex | Respiratory Rate (per minute) | Oxygen Saturation (%) | Any Supplemental Oxygen | Temp. | Systolic BP (mm Hg) | Heart Rate (per min) | NEWS |
| --- | --- | --- | --- | --- | --- | --- | --- | --- | --- |
| 1 | 50-60 | Male | 24 | 93 | yes | 98.0 | 110 | 76 | 6 |
| 2 | 40-50 | Male | 21 | 93 | yes | 98.6 | 138 | 88 | 5 |
| 3 | 40-50 | Male | 27 | 95 | yes | 97.6 | 136 | 87 | 5 |
| 4 | 50-60 | Male | 28 | 91 | yes | 98.0 | 159 | 77 | 7 |
| 5 | 30-40 | Male | 22 | 91 | yes | 98.0 | 126 | 98 | 5 |
| 6 | 50-60 | Male | 22 | 95 | yes | 98.8 | 105 | 80 | 5 |
| 7 | 50-60 | Male | 24 | 93 | yes | 98.4 | 110 | 104 | 7 |
| 8 | 40-50 | Male | 22 | 93 | yes | 98.4 | 103 | 88 | 6 |
| 9 | 50-60 | Male | 22 | 92 | yes | 98.2 | 123 | 96 | 6 |
| 10 | 60-70 | Male | 22 | 95 | yes | 98.8 | 120 | 98 | 5 |

Abbreviations DM = diabetes mellitus; HT = hypertension; NEWS = national early warning score

LDRT was delivered after a mean of 3 days (range 1-9 days) of indoor admission **(Table 3)**. Two patients (patient no. 3 and 8) received LDRT after 6 and 9 days of hospitalization respectively as they were having mild symptoms initially. All patients completed the prescribed LDRT. Average time taken for LDRT (from entering to exiting treatment room) was 11 minutes (8-34 minutes). One patient took unexpectedly longer time during treatment (34 minutes) due to technical problem in switching on the machine. Another patient took 19 minutes since the radiation therapy technologist had difficulty in starting the treatment due to fogging inside wall of face shield. No patient required RT interruption due to deterioration of vitals or oxygen saturation.

**Table 3:** Clinical Response post LDRT.

| Patient Serial No. | Hospital stay (days) | Interval between day of hospitalization and LDRT (days) | Day-0 NEWS (on the day of LDRT) | Day-3 NEWS | Day-7 NEWS | Day-14 NEWS | Day-30 clinical status |
| --- | --- | --- | --- | --- | --- | --- | --- |
| 1 | 10 | 3 | 6 | 1 | 0 | 0 | Alive |
| 2 | 24 | 2 | 5 | 0 | 0 | 0 | Alive |
| 3 | 18 | 9 | 5 | 0 | 0 | 0 | Alive |
| 4 | 15 | 1 | 7 | 2 | 1 | 0 | Alive |
| 5 | 13 | 1 | 5 | 0 | 0 | 0 | Alive |
| 6 | 24 | 2 | 5 | 6 | 9 | - | Dead |
| 7 | 15 | 2 | 7 | 4 | 0 | 0 | Alive |
| 8 | 12 | 6 | 6 | 1 | 0 | 0 | Alive |
| 9 | 14 | 2 | 6 | 3 | 0 | 0 | Alive |
| 10 | 12 | 2 | 5 | 1 | 0 | 0 | Alive |

Abbreviations: LDRT = low dose radiation therapy; NEWS = national early warning score; ICU = intensive care unit.

**Table 3** shows the progress in NEWS post LDRT. One patient, showed clinical deterioration and had to be intubated. He finally succumbed of ARDS on day 24. Rest 9 patients had complete clinical response and finally discharged from the hospital after their COVID test was negative. The average hospital stay of the cured patients was 15 days (range 10-24 days). **Fig. 1** depicts the speed of clinical recovery. Most patients achieved a NEWS of 0 by Day-7. Day-30 clinical status was determined by communicating the patients or relatives on phone. All 9 patients discharged from hospital were alive. No patients showed the signs of acute radiation toxicity.

**Fig 1:**
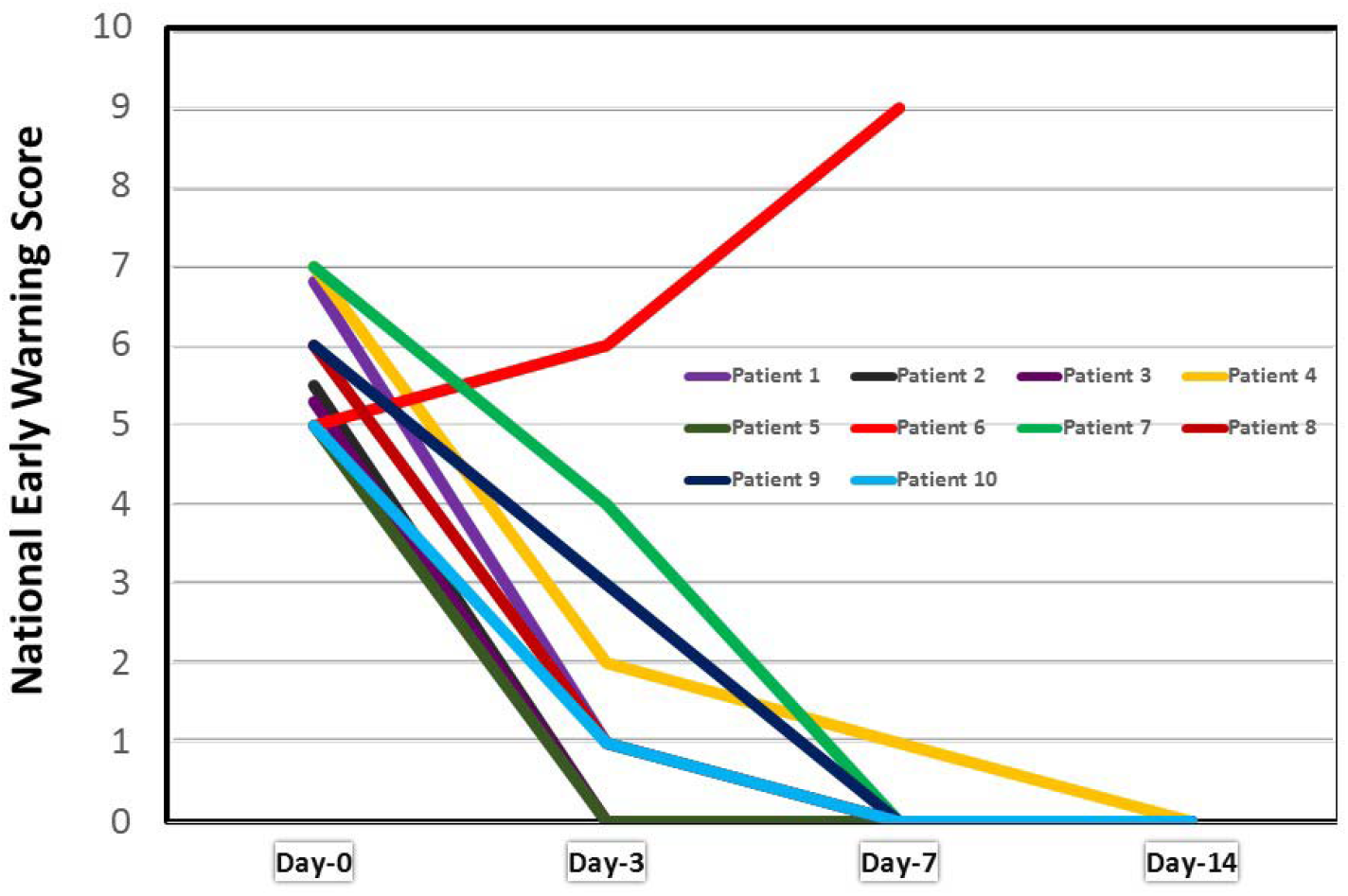
Graph showing post LDRT response as per National Early Warning Score.

## DISCUSSION

The results of our study have proven the feasibility of using LDRT for treatment of COVID-19 patients with moderate to severe risk disease. All patients completed the prescribed treatment without any hurdles. None of the radiation therapy team members involved in LDRT of these patients acquired COVID-19 infection. We used simple, open field technique for RT to minimize the radiation planning and delivery time. No patient showed any signs of acute toxicity and therefore it is clinically safe.

In terms of clinical improvement, we observed 90% clinical response rate in our study which This suggests that LDRT is clinically efficacious in COVID-19 patients with moderate to severe illnes.

Only one patient, who had associated co-morbidity (HT) did not respond to LDRT and died of ARDS. It is well known that the patients with co-morbidities have higher risk of mortality. The other 2 patients in our series (patient no. 5 and 7) who had co-morbidities had good response and recovered within a week and discharged in less than 2 weeks of LDRT. Overall results of our study have been very encouraging and justifies the conduction of future randomized controlled trial with larger sample size. Besides clinical efficacy, LDRT also helps in averting the need of ICU admission. ICU care consumes lot of manpower resources and cost which are constrained in developing countries like India having high incidence of COVID-19.

The initial report using X-rays to treat patients with pneumonia was in 1907 by Edsall and Pemberton **(9)**. Subsequent reports till 1940s suggested that LDRT was successful in decreasing the mortality rate in untreated patients from about 30% to 5-10%. **(10-13)** In 1943, Oppenheimer **(13)** reported the results of LDRT using 50 rontgen in 56 patients of viral pneumonia and observed cure rate of 80%. During that time, there was no concept of lung correction factor and therefore the actual dose delivered to lungs in Oppenheimer’s study was more than 50 rontgen. He further observed that if radiation was delivered in the first week of disease phase, the cure rate was 100% as compared to 50% when delivered after 2 weeks. Despite good clinical results reported in these trials **(10-13)**, LDRT slowly vanished after the arrival of penicillin. Kirkby and Mackenzie first suggested the use of LDRT in current COVID-19. **(16)**

Recently, two studies **(14, 15)**, to the best of our knowledge, have been published reporting the use of LDRT for COVID-19 and ours is the third one being reported here. **Table 4** shows the comparison of these two studies with our present study. Our study has the largest sample size (10 vs 5 vs 5 patients). Both trials **(14, 15)** had higher median age and co-morbidities as compared to ours. LDRT dose was highest in the study by Hess at al. **(15)** Though it is difficult to compare clinical outcome due to small sample sizes in all three studies **(Table 4)**, the recovery rate was relatively higher in our study (90% vs 80% vs 80%). This is probably because our patients had relatively less severe disease and lower frequency of co-morbidities. We believe LDRT is most effective in averting the cytokine storm and therefore may be used before the cytokine storm has set in. Therefore our study was designed to enroll patients with moderate to severe illness.

**Table 4:**
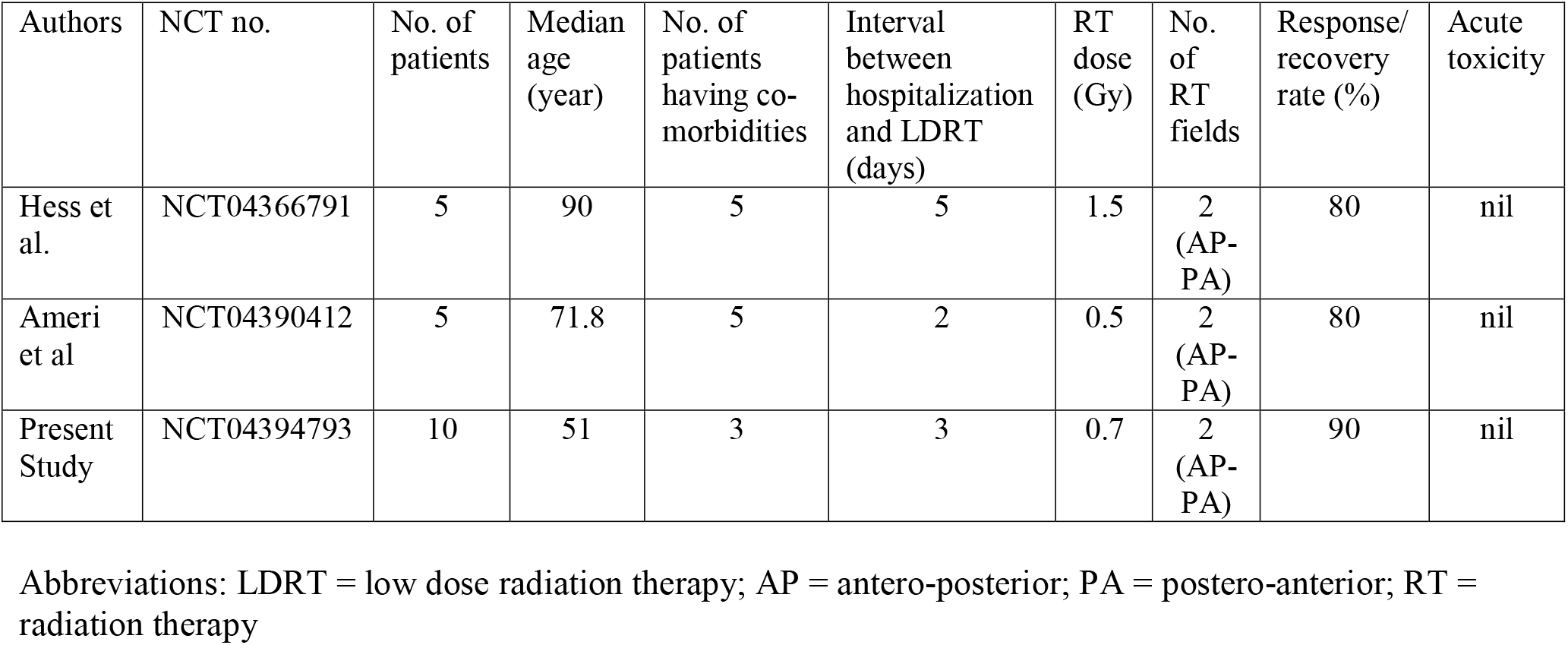
Studies published so far using LDRT for Covid-19.

Castillo et al. **(17)** have recently published a report of 64 year old patient treated with LDRT. They adopted CT based planning prescribing a dose of 1.0 Gy with VMAT technique. Clinical target volume (CTV) consisting of whole lung and (OAR) were contoured. A circumferential 5-mm and craniocaudal 10-mm PTV expansion was created. Procedure duration consisted of 30 minutes planning and about 13 minutes of treatment delivery (including cone beam CT scan performance). The patient showed improvement after 3 days of LDRT and shifted out of ICU after 6 days. We do not encourage CT based planning considering negligible dose to OAR due to low prescription dose.

Although it is hypothesized that LDRT potentially mitigate the COVID-19 pneumonitis by inducing an anti-inflammatory effect; animal, human and in vitro studies indicate that LDRT may have the potential to control bacterial pneumonia. **(18)** Therefore, LDRT may also be capable of reducing bacterial co-infections in patients with COVID-19. Additionally, LDRT might prevent accelerated viral drug-related mutation thus potentially improving the immune response by means of the enhanced RNA damage compared to antiviral therapy. **(18, 19)**

Whenever RT is employed for benign conditions, concerns are expressed about risk of radiation induced carcinogenesis. It is often ignored that even without radiation exposure; a healthy human being does carry a certain amount of life-time risk of developing cancer. The excess absolute risk (EAR) induced by radiation exposure is determined by the below formula proposed by Preston considering the β = 10, θ = − 0.05 and γ = 1.

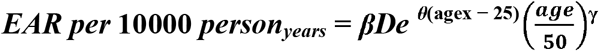

According to this formula, the increase in EAR is about 0.4% for 0.5 Gy and 1.2% for 1.5 Gy over a 20 year period. The LDRT involves only thoracic organs as OAR rather than whole body exposure thus further minimizing the risk. This EAR is negligible considering the potential benefit of LDRT in the current pandemic.

Our study has several limitations including small sample size and response assessment without radiological or laboratory investigations. We wanted to keep the study design simple and convenient for smooth conduction considering the fear of radiation exposure amongst the patient population and also the panic in radiation therapy team which is primarily a non-COVID team. There are several ongoing trials (ClinicalTrials.gov Identifier: NCT04427566, NCT04572412, NCT04377477, NCT04493294 and NCT04393948) with primary outcome assessment based on clinical parameters. **(21)**

In conclusion, the results of our pilot study suggest that LDRT is feasible and clinically effective in COVID-19 patients having moderate to severe disease. Based on the encouraging results of two recently published trials **(14, 15)** and our study, it is justified to conduct large randomized controlled trials to establish the clinical efficacy of LDRT to reduce the COVID-19 mortality.

## Data Availability

Data will be made available by authors on request.

## Acknowledgments

Authors express their gratitude to Dr KP Haresh, Dr Surendra Saini, Dr Rambha Pandey, all the residents doctors, medical physicists and therapists of the radiation oncology department for their support.

## Interpretation

- A low score (NEWS 1–4) should prompt assessment by a competent registered nurse who should decide if a change to frequency of clinical monitoring or an escalation of clinical care is required.
- A medium score (i.e. NEWS of 5–6 or a RED score) should prompt an urgent review by a clinician skilled with competencies in the assessment of acute illness – usually a ward-based doctor or acute team nurse, who should consider whether escalation of care to a team with critical-care skills is required (ie critical care outreach team).
  - A RED score refers to an extreme variation in a single physiological parameter (i.e., a score of 3 on the NEWS chart in any one physiological parameter, colored RED to aid identification; e.g., heart rate
- A high score (NEWS ≥7) should prompt emergency assessment by a clinical team/critical care outreach team with critical-care competencies and usually transfer of the patient to a higher dependency care area.

